# Unexposed populations and potential COVID-19 burden in European countries as of 21st November 2021

**DOI:** 10.1101/2021.11.10.21266166

**Authors:** Lloyd A C Chapman, Rosanna C Barnard, Timothy W Russell, Sam Abbott, Kevin Van Zandvoort, Nicholas G Davies, Adam J Kucharski

## Abstract

We estimate the potential remaining COVID-19 burden in 19 European countries by estimating the proportion of each country’s population that has acquired immunity to severe disease through infection or vaccination. Our results suggest that many European countries could still face a substantial burden of hospitalisations and deaths, particularly those with lower vaccination coverage, less historical transmission, and/or older populations. Continued non-pharmaceutical interventions and efforts to achieve high vaccination coverage are required in these countries to limit severe COVID-19 outcomes.

---

Although many European countries have experienced high burdens of COVID-19 and have managed to achieve reasonably high vaccination coverage, there are still a large number of people unvaccinated in most countries and it is unclear how many of these individuals have not yet been infected and lack immunity to the virus. Where the number of these unexposed individuals is large there is the potential for a considerable remaining burden of hospitalisations and deaths, so it is crucial to quantify how large this remaining susceptible group could be. We therefore estimate the number of unvaccinated and unexposed individuals in 19 European countries as of 21st November 2021 and introduce a metric to quantify and compare the potential remaining COVID-19 burden across countries, accounting for differences in historical burden, vaccine efficacy and population age structure.

## Determinants of remaining burden

The potential remaining COVID-19 burden in a country depends on several factors, but the key determinants are: (i) the proportion of each age group in the population with naturally-acquired immunity, (ii) the proportion of each age group with vaccine-induced immunity, and (iii) the age structure of the population, which determines the population-level risk of severe COVID-19 outcomes (hospitalisation/death). Estimating the level of naturally-acquired immunity in the population from case incidence data is difficult due to variation in reporting levels over the course of the pandemic and across countries. With reliably reported death time series and high-quality estimates of the infection fatality risk (IFR), infection time series can in principle be inferred by deconvolving the death time series and scaling it by the inverse of the IFR (1,2). However, since vaccination reduces the risk of death given SARS-CoV-2 infection and most countries have followed an age-based vaccine rollout, the IFR for COVID-19 has varied asynchronously between age groups over time, and this needs to be accounted for in the calculation. We therefore use age-stratified death (3,4) and vaccination (5) data to infer age-stratified infection time series, accounting for the impact of vaccination on the IFR for each age group over time.

## Vaccination and the infection fatality risk

COVID-19 vaccines have been shown to offer strong protection against disease, hospitalisation and death, but weaker protection against infection (6,7). Their efficacy against different outcomes also varies according to vaccine type, dose and virus variant (6). Thus, as vaccination coverage has increased (Figure 1A), the overall IFR has decreased, but in a way that depends on the speed and composition of the vaccine rollout (by age and vaccine product) and the changing proportions of different SARS-CoV-2 variants. We account for the varying impacts of vaccination on different disease outcomes by estimating the overall time-varying IFR for each age group for each country as a weighted average of the IFR for unvaccinated individuals and that for vaccinated individuals (accounting for protection against infection from vaccination), averaging the efficacies of different vaccine types and doses against different outcomes for the different circulating variants according to their relative proportions in each country (see Supplement). This gives the country-level population-weighted IFRs shown in Figure 1B. The pattern of decrease in the IFR is similar for most countries in Europe, except England and Romania, where respectively earlier and slower vaccine rollouts have led to earlier and slower declines, and the pre-vaccination IFR was higher (close to 1% vs an average of 0.8%) for countries with older populations, such as Italy and Portugal.

**Figure 1.**
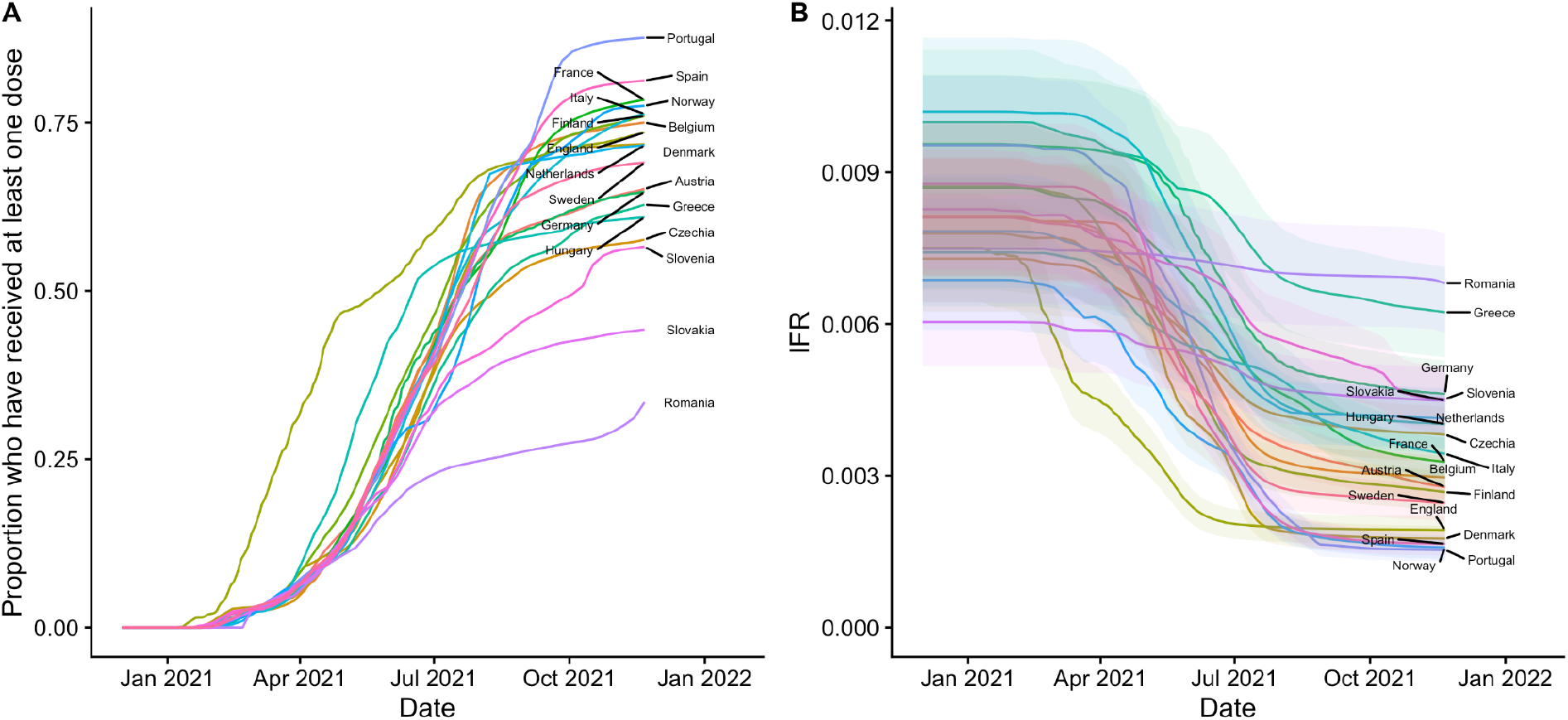
(A) Overall first-dose vaccination coverage over time since 1st December 2020 in 19 European countries and (B) corresponding population-weighted average infection fatality risk (IFR) over time. Shaded bands in (B) show 95% credible intervals based on a truncated normal approximation to posterior distribution of IFR in (8). See Supplementary Figure S1 for vaccination coverage and IFR over time by age group for each country.

## Historical burden and naturally-acquired immunity

Using the age- and time-dependent IFR for each country (Supplementary Figure S1B) we infer age-stratified infection time series from age-stratified death time series (9,10) for each country (Figure 2A), and from these calculate the cumulative proportion of (unvaccinated and vaccinated) individuals who have been infected and therefore have some degree of immunity. We make the simplifying assumption that naturally-acquired and vaccine-induced immunity do not wane, given the relatively slow waning of vaccine-induced immunity against hospitalisation and death suggested by current evidence (11) and evidence that reinfections are relatively rare and generally milder than first infections (12), and assume complete cross-protection between different variants, given limited observation of immune escape for the Alpha and Delta variants (13). We find considerable variation in the estimated proportion of the population with infection-acquired immunity between countries, ranging from 3% (95% credible interval (CI) 2-5%) in Norway over the whole population to 74% (95% CI 66-80%) in Romania.

**Figure 2.**
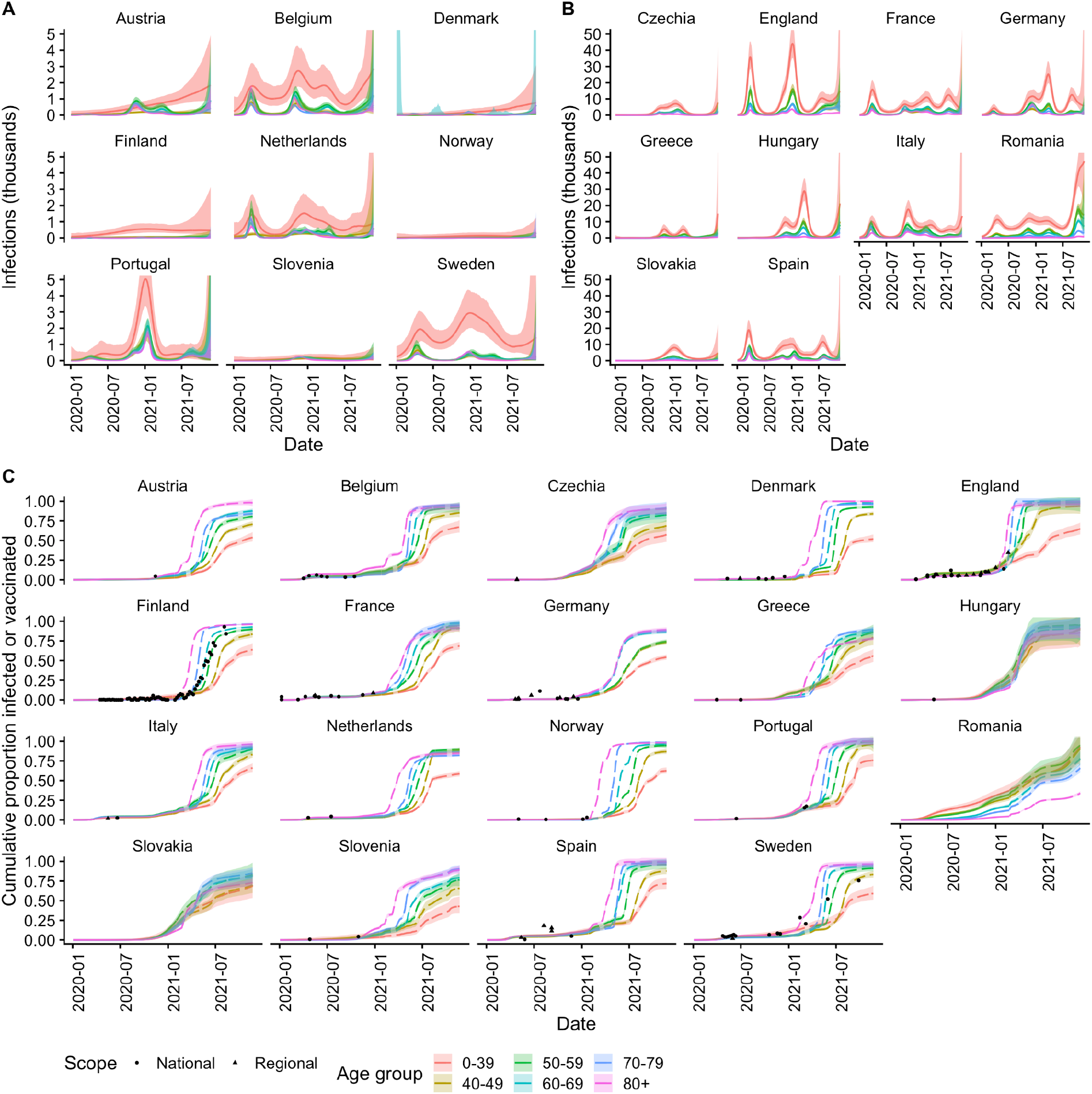
(A)-(B) Age-stratified infection time series inferred from age-stratified death time series (Supplementary Figure S2) for countries with (A) <2 million and (B) ≥2 million estimated cumulative infections. (C) Cumulative proportion infected or vaccinated against seroprevalence estimates from serological surveys in the SeroTracker database. (14). Solid lines and shaded bands show maximum a posteriori/median estimates (in (A)-(B) and (C) respectively) and 95% credible intervals for inferred infections; dashed lines in (B) show where vaccination has contributed to immunity. Points in (C) show central estimates from seroprevalence surveys plotted at the mid-point of the survey period, with geographic scope as shown in the legend. Supplementary Figure S3 shows the estimated cumulative proportion infected or vaccinated for England against modelled age-stratified seroprevalence estimates from the UK Office for National Statistics.

## Vaccine-induced immunity and breakthrough infections

To calculate the proportion of the population with immunity acquired through infection or vaccination, *or both*, it is necessary to account for some infections being breakthrough infections, i.e. among vaccinees, and some vaccinations being given to previously infected individuals, to avoid double counting these individuals when tallying the overall number with immunity. We do this by solving a difference equation model for the movement of individuals between susceptible, vaccinated and infected states over time in each age group with the backcalculated infection estimates as the input for the number of infections occurring at each time step (see Supplement). The relative proportions of infections at each time step coming from susceptible and vaccinated individuals are assumed to be proportional to their susceptibility-weighted relative prevalences, and the proportion of vaccinations that go to susceptible individuals is assumed to be equal to the proportion of individuals still to be vaccinated who are susceptible. This approach suggests that in many countries the proportion of individuals in older age groups (≥50yrs) who are both unvaccinated and unexposed is small (<10%) (Figures 2C and 3A), and that the majority of unexposed unvaccinated individuals are in younger age groups (<40yrs), with the overall proportion of unvaccinated and unexposed individuals varying from 5% (95% CI 1-9%) in Hungary to 37% (95% CI 33-39%) in Slovenia (Supplementary Table S4).

## Ethical statement

Ethical approval was not necessary for this modelling study as the analysis uses only publicly available aggregated secondary data.

### Remaining hospitalisations and deaths

Having estimated the remaining number of unvaccinated and unexposed individuals in each age group in each country, we calculate the potential remaining COVID-19 burden as the number of hospitalisations/deaths that would occur per 100,000 individuals if the entire population were to be (re-)exposed now (Figure 3). In other words, the burden if all unexposed and unvaccinated individuals were infected and a proportion of vaccinated individuals and previously infected individuals were infected dependent on the protection against infection afforded by vaccination/previous infection. This represents an upper bound on the potential burden of hospitalisations and deaths that could occur in the absence of waning of immunity and any further vaccination, as in reality not all unexposed and unvaccinated individuals would become infected in an uncontrolled epidemic (15). Nevertheless, it does not account for waning of immunity, population growth or potential emergence of immune escape variants, all of which could increase the burden of hospitalisations and deaths.

**Figure 3.**
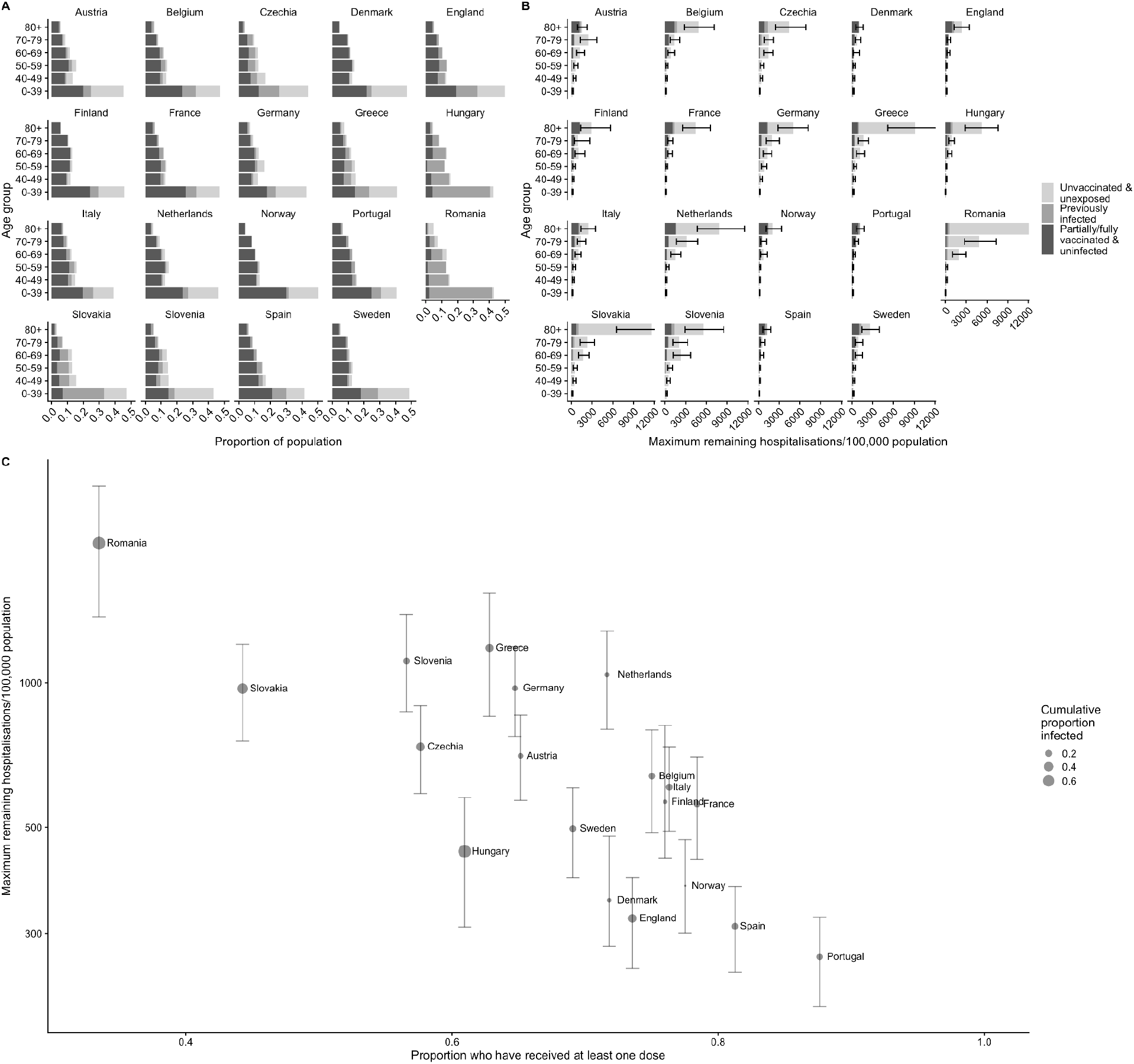
(A) Population pyramids showing the proportions of each age group in each country that are partially/fully vaccinated (and not previously infected), previously infected, and unvaccinated and unexposed. (B) Estimated maximum remaining hospitalisations per 100,000 population by age group, country and immune status (assuming no waning of immunity or emergence of immune escape variants), and (C) relationship between overall remaining hospitalisations per 100,000 population and proportion who have received at least one vaccine dose across countries. Error bars in (C) show 95% credible intervals. Note horizontal axis in (B) has been truncated to ensure differences between countries remain visible and vertical axis in (C) is on log scale. See Supplementary Figure S4 for pattern of maximum remaining deaths by age and country and its relationship with vaccination coverage, and Supplementary Figure S5 for the relationship between remaining hospitalisations and deaths and population age structure. Supplementary Figure S6A shows plot (B) without truncation of the horizontal axis.

The estimated maximum overall remaining hospitalisations per 100,000 individuals ranges widely across countries, from 270 (95% CI 210-320) in Portugal to 2000 (95% CI 1400-2600) in Romania (Figure 3C and Supplementary Table S4). This reflects the higher vaccination coverage in Portugal and the lower vaccination coverage, particularly among older individuals, in Romania than other countries (Figure 3A). In absolute terms, the estimated maximum number of remaining hospitalisations ranges from 20,000 (95% CI 16,000-28,000) for Denmark to 820,000 (95% CI 650,000-1,000,000) for Germany. The comparative pattern of maximum remaining COVID-19 deaths across countries is similar (Supplementary Figure S4), given the strong influence of age on the IFR as well as the IHR (8,16), with the burden of deaths per 100,000 people ranging from 50 (95% CI 44-58) for Portugal to 540 (95% CI 450-630) for Romania, and absolute numbers of deaths ranging from 2,900 (95% CI 2,500-3,900) for Denmark to 170,000 (95% CI 150,000-190,000) for Germany. Aside from Romania, where vaccination coverage is low, countries with a combination of lower vaccination coverage among older age groups, relatively low prior exposure and older populations (Germany, Greece, the Netherlands, Slovenia) have the highest maximum remaining burdens (Figure 3C and Supplementary Figure S5), as they have the potential to experience much higher numbers of hospitalisations and deaths among the elderly than countries with younger populations and high coverage in older age groups.

## Discussion

Our results suggest that the potential remaining burden of COVID-19 hospitalisations and deaths across the 19 European countries considered is substantial, amounting to over 3 million hospitalisations (95% CI 2.4-3.8 million) and 600,000 deaths (95% CI 550,000-750,000), but that it varies considerably between countries, with countries that have experienced less transmission so far, have lower vaccination coverage and/or have older populations having much higher potential remaining burdens. The main driver of the potential remaining burden according to our analysis is vaccination coverage achieved so far, with the proportion of the population infected to date and the age structure of the population having lesser effects, as the speed of the vaccine rollout has meant that vaccine-induced immunity dominates naturally-acquired immunity in most countries. Countries that appear to be at particular risk of high burden include Romania (due to its low vaccination coverage), Greece, Slovenia, the Netherlands, Slovakia and Germany. Portugal, Spain, England, Denmark and Norway meanwhile appear to have lower risk due to much higher vaccination coverage.

This analysis has a number of limitations (see Supplement for a full discussion). Our estimates only provide an upper bound on the burden that could theoretically occur in the short-term from a complete lifting of non-pharmaceutical interventions and return to pre-pandemic contact levels with no further vaccination, not an indication of the burden that will actually occur in each country. We do not consider longer-term burden, as it will depend on the impact of waning immunity on severe disease, which is still being determined (11), and would also be influenced by susceptible individuals being replenished through births. We do not account for waning of immunity against infection or severe outcomes, the rollout of boosters or the potential emergence of new variants. Accounting for waning of immunity would increase estimates or potential remaining burden, while boosters would likely decrease them, given evidence of increased neutralising antibody titres and protection following booster administration (17–19). The emergence of new variants, such as Omicron, which show immune escape properties (20), could significantly increase future burden and reduce the impact of the proportion vaccinated/previously infected, depending on the extent to which they can evade immunity against hospitalisation and death from vaccination/previous infection.

## Conclusion

Continued non-pharmaceutical interventions and efforts to achieve high vaccination coverage are required in the short term in European countries to limit severe COVID-19 outcomes. Exactly how transmission dynamics evolve in different countries in the longer term will depend on local policies, and this will be explored through more detailed modelling building on this analysis.

## Data Availability

All data and code used in this manuscript are available online at https://github.com/LloydChapman/covid_remaining_burden and https://doi.org/10.5281/zenodo.5772163 or at the links below.

https://osf.io/mpwjq/

https://dc-covid.site.ined.fr/en/

https://www.ecdc.europa.eu/en/covid-19/data

https://coronavirus.data.gov.uk/details/download

https://covid19.sanger.ac.uk/downloads

https://serotracker.com/

https://doi.org/10.5281/zenodo.5772163

## Acknowledgements

We gratefully acknowledge the GISAID initiative and all the authors from the originating laboratories where genetic sequence data were generated for sharing such data through GISAID, and enabling the estimation of SARS-CoV-2 variant proportions in this work.

LACC is grateful to Marinella Capriati and to Carl Pearson of the London School of Hygiene and Tropical Medicine for helpful discussions, and to Nik Kolb of IQVIA and Florian Schumann of Die Zeit for help obtaining and interpreting age-stratified vaccination data for Germany.

## Authors’ contributions

LACC and AJK conceptualised the study and devised the methodology. LACC carried out the analysis and wrote the first draft of the manuscript. RCB compiled the information on vaccine efficacies. RCB, TWR, SA, KvZ and NGD contributed to the analysis and interpretation of the data and presentation of figures, and reviewed and edited the manuscript. All authors approved the final version of the manuscript for submission.

