## Supplement for "Unexposed populations and potential COVID-19 burden in European countries as of 21st November 2021"

#### Methods

##### Data and data processing

###### Deaths

We use country-level age-stratified data on COVID-19 deaths in 19 European countries from 2nd January 2020 to 21st November 2021 from the COVerAGE database (1) – a global demographic database of COVID-19 cases and deaths – which is freely available from the Open Science Framework (2), and from the Institut National d'Études Démographiques (INED) database (3). The COVerAGE data is pre-cleaned and processed to harmonise metrics and age groups (into 10-year intervals up to age 100yrs and then 100-104yrs) across countries (see (2) for details). However, both datasets have periods of missing data and negative counts in the data due to downward corrections of death totals by country health ministries, and the COVerAGE dataset has some errors introduced through the pre-processing. We first clean the data by:

i. Correcting typographical errors that occurred in the entry of the raw data.
ii. Removing values for dates where there is a clear error, e.g. the cumulative death total is lower than for the previous date or a change in the age groups reported in the raw data has led to an error in the age group reaggregation.

We then calculate the cumulative deaths across all ages for each country and merge the dataset with World Health Organization (WHO) country-level data on cumulative deaths, downloaded and cleaned using the `covidregionaldata` R package (4). We "fill in" periods of missing data at the start of the pandemic and longer periods of missing data (>1 month) in the COVerAGE data for Finland by scaling the cumulative number of deaths in each age group at the end of the period of missing data by the distribution of total deaths over time in the WHO data for that period. We also extrapolate the death time series in recent days/weeks where the age-stratified data for some countries is not fully up-to-date by multiplying the cumulative numbers of deaths in each age group at the most recent time point available by the distribution of total deaths over time since that point in the WHO data. We reaggregate the deaths into the age groups used in the analysis (0-39, 40-49, 50-59, 60-69, 70-79, 80+yrs), and linearly interpolate cumulative deaths for each age group between successive dates where there are short periods of missing values. Daily numbers of deaths are then calculated by differencing the cumulative death time series.

###### Vaccinations

We use age-stratified data on weekly numbers of vaccinations with different vaccine types in EU countries from 14th December 2020 to 21st November 2021 from the European Centre for Disease Prevention and Control (ECDC) (5) for European Union countries and aggregated age-stratified data on daily numbers of vaccinations with different vaccine types from 8th December to 21st November 2021 for England from the UK government's COVID-19 dashboard (6). For the ECDC data, we convert weekly numbers of vaccinations into daily numbers of vaccinations by assuming equal numbers of vaccinations occurred on each day of a given week. We categorise the different vaccines into two groups – (A) viral vector and inactivated virus vaccines (AstraZeneca, Janssen, Sputnik and Beijing CNBG), and (B) mRNA vaccines (Pfizer and Moderna) – based on higher efficacies for mRNA vaccines than viral vector and inactivated virus vaccines but similar efficacies within these groups, and these being the vaccine types that have been used in European countries. First and second doses of unknown vaccine type are assigned a vaccine type according to the average proportion of each type used for first and second doses for the given age group during that week in other countries. Vaccinations with an unknown age group constitute only 0.008% of all vaccinations (and less than 0.3% of total vaccinations in any one country) so are disregarded. The breakdown of vaccinations by age is missing from the ECDC data for the Netherlands, so we scale the total number of vaccinations by the mean proportion of vaccinations in each age group in each rollout week across countries with age-stratified data (weighted by the relative population proportions of the age groups in each country) to estimate the vaccinations in each age group. Vaccination data for Germany is only available in broader age groups (<18, 19-59, 60+yrs) than we use in the analysis (7). We therefore use data from the Robert Koch Institute's COVIMO phone survey of vaccine uptake in German citizens (8) (Supplementary Table S2) to estimate the relative coverages in the finer age groups within each of broader age groups, and scale the total numbers of vaccinations over time in the broad age groups by these relative coverages. Whilst this does not fully account for the initial age prioritisation of the vaccine rollout in Germany (Supplementary Figure S1A), it gives more accurate estimates of the current vaccine coverage in the eldest age groups than the method used for the Netherlands, which underestimates coverage in the eldest age groups in Germany. We assume fixed delays of 28 days and 14 days for development of protection following first and second doses, corresponding to the time points at which efficacy was measured in clinical trials (Supplementary Table S1).

#### Variants

We use data on frequencies of SARS-CoV-2 variants among sequenced infections in each country from the Global Initiative on Sharing Avian Influenza Data (GISAID) EpiCoV database (9) and The European Surveillance System (TESSy) (10) (collated by the ECDC (11)), and from the COVID-19 Genomics UK (COG-UK) consortium Lineages database (12). We classify variants into three groups – "Other" for pre-existing (pre-Alpha) and other variants, "Alpha" (B.1.1.7), and "Delta" (all B.1.617.2 variants) – as at the time of writing these are the main variants that have emerged and circulated in European countries.

#### Vaccine efficacies

We use estimates of overall vaccine efficacies against different outcomes – infection ($i$), disease ($d$), hospitalisation ($h$), and death ($m$) – for different doses of different vaccines and different variants from various efficacy studies to calculate the efficacies required for our model: those against infection, against disease given infection, against hospitalisation given disease, and against death given disease. We assume that the efficacies against each outcome of the different vaccines are the same for “Other” variants and the Alpha variant. If $e_i$ is the vaccine efficacy against infection and $oe_o$ is the overall efficacy against outcome $o \in \{d, h, m\}$, then we calculate the efficacies against disease given infection, hospitalisation given disease, and death given disease, $e_d$, $e_h$, and $e_m$ respectively, for each dose and variant as:

$$e_d = \frac{oe_d - e_i}{1 - e_i}$$

$$e_h = \frac{oe_h - oe_d}{(1 - e_i)(1 - e_d)}$$

$$e_m = \frac{oe_m - oe_d}{(1 - e_i)(1 - e_d)}$$

Supplementary Table S1 shows the values of the vaccine efficacies we use in the model and their sources.

#### Infection fatality risk

For the age-specific SARS-CoV-2 infection fatality risk (IFR) in unvaccinated individuals used in the analysis, we use the ensemble IFR estimate from (13), which combined data on age-stratified COVID-19 deaths up to 1st September 2020 for 45 countries with data from 22 national seroprevalence surveys to estimate the age-specific IFR. COVID-19 deaths were split by long-term care (LTC) residency status in (13) to account for differences in nursing home burdens across countries, so the estimated IFR represents the IFR in non-LTC residents. We incorporate uncertainty in the IFR into our estimates by assuming that the posterior distribution for the IFR for each age group $a$, $IFR_{u,a}$, is approximately truncated normal, with mean equal to the median estimate, $\widetilde{IFR}_{u,a}$, standard deviation $\sigma_a$ determined from the lower bound of the 95% credible interval reported in (13), and truncation at 0:

$$IFR_{u,a} \sim TN(\widetilde{IFR}_{u,a}, \sigma_a^2, 0, \infty). \quad (1)$$

#### Population age distributions

We use the UN World Population Prospects 2020 population estimates by single-year age from 0-100yrs (14) for the population age distributions of each country. We make the simplifying assumption that population sizes and age distributions have remained constant over the course of the pandemic.

#### Backcalculation of infections from age-stratified death data

The number of deaths in a particular age group $a$ on a particular day $t$, $D_{a,t}$, is related to the number of infections on preceding days, $Inf_{a,t-s}$ $(0 \leq s < t)$, by the convolution of the infection counts with the distribution of the infection-to-death delay, $d_{Death}$, and scaling with the IFR for age group $a$ on those days, $IFR_{a,t-s}$, i.e.:

$$D_{a,t} = \sum_{s=0}^{t} IFR_{a,t-s} Inf_{a,t-s} p_s$$

where $p_s = P(d_{Death} = s)$ is the probability of a delay of $s$ days between infection and death. Note that here, unlike in other studies (15–17), the IFR depends on time of infection, because the IFR changes as vaccine coverage increases.

We use the convolution of the latent period distribution (the time between infection and start of infectiousness), $d_E$, and the time from start of infectiousness to death, $d_{Inf,Death}$, for the infection-to-death delay, $d_{Death} = d_E * d_{Inf,Death}$. We use $d_E \sim Gamma(2.5, 1)$ (mean 2.5 days, standard deviation 1.6 days) and $d_{Inf,Death} \sim Gamma(2.2, 6.8)$ (mean 15 days, standard deviation 10 days) (18), where $Gamma(k, \theta)$ denotes the gamma distribution with shape parameter $k$ and scale parameter θ. Since the data is in discrete time with time steps of a day, we discretise these distributions by integrating the probability density functions over the half day either side of the start of each day, and use the discrete convolution to find the infection-to-death delay distribution. This gives a distribution with a mean of 17.4 days and a standard deviation of 9.9 days.

##### Calculation of time-varying infection fatality risk

A number of factors are likely to have affected the SARS-CoV-2 IFR over the course of the pandemic, including surges in demands for hospital beds, improvements in quality of care for hospitalised COVID-19 patients, differences in variant severity, and vaccination. For the sake of simplicity, and because we believe it is likely to have had the largest influence, we consider only the impact of vaccination on the IFR in this analysis. We model the time-varying IFR as a weighted average of the IFR for unvaccinated individuals and that for vaccinated individuals. Specifically, we want the IFR to reflect the expected split in deaths between unvaccinated individuals and vaccinated individuals over time based on the efficacies of the vaccine against infection and against death given infection:

$$\begin{aligned} D_{a,t} &= \sum_{s=0}^{t} IFR_{a,t-s} Inf_{a,t-s} p_s \\ &= D_{u,a,t} + D_{v,a,t} \end{aligned}$$

$$= \sum_{s=0}^{t} (IFR_{u,a} Inf_{u,a,t-s} + IFR_{v,a} Inf_{v,a,t-s}) p_s,$$

where subscripts $u$ and $v$ represent quantities for unvaccinated and vaccinated individuals respectively and $Inf_{a,t} = Inf_{u,a,t} + Inf_{v,a,t}$, i.e. we want:

$$IFR_{a,t} = IFR_{u,a} \frac{Inf_{u,a,t}}{Inf_{a,t}} + IFR_{v,a} \frac{Inf_{v,a,t}}{Inf_{a,t}}$$
$$= IFR_{u,a} (1 - \frac{Inf_{v,a,t}}{Inf_{a,t}}) + IFR_{v,a} \frac{Inf_{v,a,t}}{Inf_{a,t}}.$$

Vaccine-induced immunity against hospitalisation and death wanes relatively slowly (19) and reinfections appear to be relatively rare and generally associated with milder symptoms (20), so the proportion of deaths that are from reinfections is likely to be small. This, coupled with the fact that vaccine rollout occurred sufficiently early in most European countries that the number of vaccinated individuals dominates the number of previously infected individuals, and vaccine efficacy against infection is relatively high, means that the proportion of infections in vaccinated individuals at time $t$, $Inf_{v,a,t}/Inf_{a,t}$, can be approximated as

$$\frac{Inf_{v,a,t}}{Inf_{a,t}} \approx \frac{(1-e_{i,t})c_{a,t}}{1-e_{i,t}c_{a,t}}, \quad (2)$$

where $e_{i,t}$ is the efficacy of the vaccine against infection at time $t$, and $c_{a,t}$ is the vaccine coverage in age group $a$ at time $t$ (21). The IFR for vaccinated individuals is given by:

$$IFR_{v,a} = P(death|infection, vaccination) = (1 - e_{d,t})(1 - e_{m,t}) IFR_{u,a},$$

where $e_{d,t}$ is the vaccine efficacy against disease given infection and $e_{m,t}$ is the vaccine efficacy against death given disease. Thus, the time-varying average IFR can be approximated as:

$$IFR_{a,t} \approx (1 - \frac{(1-e_{i,t})c_{a,t}}{1-e_{i,t}c_{a,t}} + \frac{(1-e_{i,t})c_{a,t}}{1-e_{i,t}c_{a,t}} (1 - e_{d,t})(1 - e_{m,t})) IFR_{u,a} \quad (3)$$

We model all vaccinations together and account for differences in the vaccine efficacies against different outcomes between different vaccine types and doses and virus variants by treating the efficacies as time dependent and averaging them over the relative proportions of the different vaccine types and doses and circulating variants in each country over time. I.e., if $e_{o,t}$ is the efficacy against outcome $o$ ($o \in \{i, d, m\}$) at time $t$, then:

$$e_{o,t} = \sum_{v} \sum_{j} p_{v,j,t} \sum_{k} p_{k,t} e_{o,v,j,k}$$

where $p_{v,j,t}$ is the proportion of vaccinees who have been vaccinated with dose $j$ of vaccine $v$ by time $t$, $p_{k,t}$ is the relative proportion of variant $k$ circulating at time $t$, and $e_{o,v,j,k}$ is the efficacy of dose $j$ of vaccine $v$ against outcome $o$ from infection/disease with variant $k$. Currently, we only consider numbers of first and second doses ($j \in \{1, 2\}$) of each vaccine, i.e. we do not include booster doses. The relative proportions of the different variants over time are estimated as described below.

#### Estimation of variant proportions over time

We estimate the relative proportions of the "Other", Alpha, and Delta variants over time in each country by fitting multinomial regression models to the frequencies of sequenced variants of each type in GISAID data and COG-UK data, in line with several other studies (18,22,23). We test two fixed-effect multinomial models: one with just sample date as the explanatory variable and one with a two-degrees-of-freedom natural cubic spline function of sample date as the explanatory variable (18). The models are fitted using the `multinom` function in the `nnet` R package and their fit is compared using Akaike Information Criterion (AIC). The model with the lowest AIC, the cubic spline model, is selected and used to calculate the variant proportions over time for each country. The input data and estimated variant proportions over time are shown in Supplementary Figure S7.

#### Deconvolution of deaths to infections

Having calculated the time-varying IFR, we estimate the time series of infections for each age group for each country using the Robust Incidence Deconvolution Estimator (RIDE) provided in the `incidental` R package (24,25), an empirical Bayes deconvolution algorithm developed and validated for inferring SARS-CoV-2 infection time series from case/hospitalisation/death time series (18,24). RIDE uses a cubic spline model for underlying infections and a Poisson likelihood to describe observed cases/deaths. The degrees of freedom of the spline basis are chosen based on lowest AIC over a grid of pre-specified values, and a regularisation penalty is imposed on the second difference of the spline parameters to encourage smoothness, and the regularisation parameter chosen by maximising held-out log-likelihood from splitting of the data into training and validation data sets over a grid of pre-set values (see (24,25) for further details). We use a grid of the even numbers between 6 and 30 for the degrees of freedom of the cubic spline – a wider range than the default setting (6 to 20) to allow sufficient flexibility to fit the observed death time series. RIDE handles right censoring of the death time series by making multiple extrapolations of the observed death curve with a simple autoregressive random walk model that has the same autocorrelation as the observed data. The mean of the autoregressive model is determined by fitting a linear model to the tail of the observed death curve over a certain time window. We use a longer time window (28 days vs 14 days) and higher prior precision (100 vs 10) for the slope of the extrapolation than the default values in order that the inferred infection curve reflects the considerable uncertainty in the future epidemic trajectory.

We first aggregate COVID-19 deaths into the following age groups: 0-39, 40-49, 50-59, 60-69, 70-79, 80+yrs. The age grouping is chosen to ensure that there are sufficient deaths in the youngest age group to reliably infer the underlying infection time series even for countries with low numbers of deaths, since the IFR increases approximately exponentially with age (13,26) and low numbers of deaths lead to unstable estimates of infections (24,27).
We then split deaths by LTC residency status as there is substantial evidence that IFRs among LTC residents have been higher than in the same age groups outside of LTC facilities (13,28–30). We assume that all deaths in LTC facilities occurred in individuals aged 60yrs and older, and use data on the proportion of all deaths that have occurred in care homes, $p_{LTC}$, for each country from the LTCcovid platform (13,28) to estimate the proportion of deaths amongst those aged 60yrs and over that have occurred in care homes. Namely, if $p_{60+}$ is the proportion of all deaths amongst individuals 60+yrs, we calculate the number of deaths amongst those 60+yrs that occur among LTC residents in each age group $a$, $D_{a,t,LTC}$, as:

$$D_{a,t,LTC} = \frac{p_{LTC}}{p_{60+}} D_{a,t}, \qquad a \geq 60yrs,$$

and the number of non-LTC deaths amongst individuals 60+yrs, $D_{a,t,nLTC}$, as:

$$D_{a,t,nLTC} = D_{a,t} - D_{a,t,LTC}, \quad a \geq 60yrs.$$

For France the reported death data is for hospital deaths only, so we estimate the proportion of deaths among those 60+yrs as:

$$p_{60+} = p_{LTC} + (1 - p_{LTC}) \frac{\sum_t \sum_{a>60} D_{a,t,Hosp}}{\sum_t \sum_a D_{a,t,Hosp}}$$

and scale up the hospital deaths among those 60+yrs to obtain overall deaths in those 60+yrs:

$$D_{a,t} = \frac{D_{a,t,Hosp}}{1 - \frac{p_{LTC}}{p_{60+}}}, \qquad a \geq 60yrs.$$

For countries for which information on the proportion of deaths among LTC residents is not available, we use the mean proportion across countries for which LTC death data is available.

With deaths split in this way, we run RIDE on the time series of deaths for each age group under 60yrs and the separate time series for non-LTC and LTC residents for each age group over 60yrs to estimate the time series of infections that led to these deaths. We then calculate the overall number of infections by dividing by a value of the IFR for the corresponding age group drawn from the distribution in Equation (1). Following (13), we take the relative frailty factor for death from COVID-19 for LTC residents to be $\gamma = 3.8$, i.e. use a 3.8 times higher IFR for LTC residents than non-LTC residents.

From the deconvolution of the death time series, RIDE produces a maximum a posteriori estimate and 1000 posterior samples of the underlying infection time series, which we use to calculate 95% credible intervals (CIs) (as the 2.5th-97.5th percentile interval) for the number of individuals that have been infected and the remaining burden of hospitalisations and deaths.

#### Calculation of proportions in different immune states

We estimate the numbers of individuals in different infection states (susceptible, vaccinated, recovered, etc.) in each age group $a$ over time $t$ from the inferred numbers of infections $Inf_{a,t}$ (for non-LTC residents and LTC residents) using the following discrete-time difference equation model (18)

$$S_{a,t+1} = S_{a,t} - nSE_{a,t} - nSV_{a,t}$$
$$V_{a,t+1} = V_{a,t} + nSV_{a,t} - nVE_{a,t} - nVL_{a,t}$$
$$E_{a,t+1} = E_{a.t} + nSE_{a,t} + nVE_{a,t} - nEI_{P,a,t} - nEI_{S,a,t}$$
$$L_{a,t+1} = L_{a,t} + nVL_{a,t} - nLI_{S,a,t}$$
$$I_{S,a,t+1} = I_{S,a,t} + nEI_{S,a,t} + nLI_{S,a,t} - nI_SR_{a,t}$$
$$I_{P,a,t+1} = I_{P,a,t} + nEI_{P,a,t} - nI_PI_{C,a,t}$$
$$I_{C,a,t+1} = I_{C,a,t} + nI_PI_{C,a,t} - nI_CR_{a,t}$$
$$R_{a,t+1} = R_{a,t} + nI_SR_{a,t} + nI_CR_{a,t}$$
$$nSE_{a,t} = \frac{S_{a,t}}{S_{a,t}+(q_t+r_t)V_{a,t}} Inf_{a,t}$$
$$nVE_{a,t} = \frac{q_tV_{a,t}}{S_{a,t}+(q_t+r_t)V_{a,t}} Inf_{a,t}$$
$$nVL_{a,t} = \frac{r_tV_{a,t}}{S_{a,t}+(q_t+r_t)V_{a,t}} Inf_{a,t}$$
$$nEI_{P,a,t} = y_a nEI_{a,t}$$
$$nEI_{S,a,t} = (1 - y_a) nEI_{a,t}$$
$$S_{a,0} = N_a,\ V_{a,0} = 0,\ E_{a,0} = 0,\ L_{a,0} = 0,$$
$$I_{S,a,0} = 0,\ I_{P,a,0} = 0,\ I_{C,a,0} = 0,\ R_{a,0} = 0.$$

Here $X_{a,t}$ is the number of individuals in state $X$ (one of: $S$ susceptible, $V$ vaccinated, $E$ latently infected, $L$ latently infected following vaccination but protected from clinical infection, $I_S$ subclinical infection, $I_P$ preclinical infection, $I_C$ clinical infection, $R$ recovered) in age group $a$ at time $t$; $nXY_{a,t}$ is the number of individuals who move from state $X$ to state $Y$ in age group $a$ at time $t$; $q_t = (1 - e_{i,t})(1 - e_{d,t})$ and $r_t = (1 - e_{i,t})e_{d,t}$ capture the average vaccine protection against disease and infection at time $t$; $nEI_{a,t}$ is the total number of individuals leaving the preclinical infection state in age group $a$ at time $t$; $y_a$ is the age-dependent clinical fraction (31); and $N_a$ is the population of age group $a$. The numbers of individuals moving between different infection states – $nEI_{P,a,t}$ , $nEI_{S,a,t}$, $nLI_{S,a,t}$, $nI_SR_{a,t}$, $nI_PI_{C,a,t}$, $nI_CR_{a,t}$– are determined by the waiting time distributions ($d_E$, $d_L$, $d_P$, $d_C$, $d_S$) for each of the infection states ($E$, $L$, $I_P$, $I_C$, $I_S$), which are given in Supplementary Table S3.

To account for the fact that not all vaccinations are administered to susceptible individuals, we model the number of vaccinations given to susceptible individuals at time, $nSV_{a,t}$, as the total number of vaccinations $nV_{a,t}$ multiplied by the proportion of those still to be vaccinated coming from the susceptible state, i.e.:

$$nSV_{a,t} = min(\frac{S_{a,t}}{(1-c_{a,t})N_a} nV_{a,t}, S_{a,t})$$

where the minimum function is to ensure the number of susceptible individuals cannot become negative. Solving the difference equations allows us to estimate how many susceptible (unexposed and unvaccinated) individuals $S_{a,T}$, uninfected vaccinees $V_{a,T}$, and previously infected individuals $R_{a,T}$ there are currently (at time $t = T$) in each age group in each country.

### Calculation of remaining burden

The maximum remaining hospitalisations $H_{rem,a}$ and deaths $D_{rem,a}$ in each age group in each country (assuming no emergence of immune escape variants) are calculated from $S_{a,T}$, $V_{a,T}$, and $R_{a,T}$ assuming that all individuals in the population are exposed now (i.e. no further vaccinations occur) and that previous infection confers a similar degree of protection against subsequent infection, hospitalisation and death as vaccination with two doses of a type A vaccine or two doses of a type B vaccine (we use the variant-proportion-weighted average of the values in Supplementary Table 1) (32). Thus all susceptible individuals become infected and a proportion of uninfected vaccinees and previously infected individuals become infected that depends on the protection against infection afforded by vaccination/previous infection. We assume that the proportion of infections in each age group ≥60yrs that occur in LTC facilities, $p_{Inf,a,LTC}$, remains the same as it is now (as estimated from the deconvolution), and assume that the relative frailty factor for hospitalisation from COVID-19 for LTC residents is the same as that for death from COVID-19. So, using estimates of the infection hospitalisation risk $IHR_{u,a}$ (33) and infection fatality risk $IFR_{u,a}$ (13) in unvaccinated individuals from the literature, $H_{rem,a}$ and $D_{rem,a}$ are given by:

$$
\begin{aligned}
H_{rem,a} &= H_{rem,a,S} + H_{rem,a,V} + H_{rem,a,R} \\
&= (1 - p_{Inf,a,LTC} + p_{Inf,a,LTC}\gamma)(IHR_{u,a} S_{a,T} + (1 - e_{i,T}) IHR_{v,a} V_{a,T} + (1 - e_{i,T}')IHR_{r,a} R_{a,T}) \\
&= (1 - p_{Inf,a,LTC} + p_{Inf,a,LTC}\gamma)IHR_{u,a} (S_{a,T} + (1 - e_{i,T})(1 - e_{d,T})(1 - e_{h,T})V_{a,T} + (1 - e_{i,T}')(1 - e_{d,T}')(1 - e_{h,T}')R_{a,T}), \\
D_{rem,a} &= D_{rem,a,S} + D_{rem,a,V} + D_{rem,a,R} \\
&= (1 - p_{Inf,a,LTC} + p_{Inf,a,LTC}\gamma)(IFR_{u,a} S_{a,T} + (1 - e_{i,T})IFR_{v,a} V_{a,T} + (1 - e_{i,T}')IFR_{r,a} R_{a,T}) \\
&= (1 - p_{Inf,a,LTC} + p_{Inf,a,LTC}\gamma)\, IFR_{u,a} (S_{a,T} + (1 - e_{i,T})(1 - e_{d,T})(1 - e_{m,T})V_{a,T} + (1 - e_{i,T}')(1 - e_{d,T}')(1 - e_{m,T}')R_{a,T}).
\end{aligned}
$$

where

$$e_{o,T}' = \frac{1}{2}\sum_{v}\sum_{k} p_{k,T} e_{o,v,2,k}$$

is the average of the type A and type B two-dose vaccine efficacies $e_{o,v,2,k}$ against outcome $o \in \{i, d, h, m\}$ weighted according to the proportions of the currently circulating variants $p_{k,T}$. We incorporate uncertainty in the estimated IHR in unvaccinated individuals from (33) by assuming that the posterior distribution for the IHR in each age group $a$, $IHR_{u,a}$, is approximately truncated normal, with mean equal to the median estimate, $\widetilde{IHR}_{u,a}$, standard deviation $\sigma_{H,a}$ determined from the lower bound of the 95% credible interval reported in (33), and truncation at 0:

$$IHR_{u,a} \sim TN(\widetilde{IHR}_{u,a}, \sigma_{H,a}^{2}, 0, \infty).$$

We calculate 95% CIs for the remaining hospitalisations and deaths by repeating the above calculations for each of the 1000 posterior samples of the infection time series generated by the deconvolution and IFR scaling (each time with a separate draw for the IHR and the draw for the IFR used in the backcalculation), and calculating the 2.5th-97.5th percentile of the resulting distribution.

#### Comparison of inferred infections with reported cases

For comparing the inferred infection time series with time series of reported cases, we convolve the inferred infection time series with the incubation period (time from infection to symptom onset), which we take to be the convolution of the latent period ($d_E \sim Gamma(2.5, 1)$) and the preclinical infectious period ($d_P \sim Gamma(4, 0.625)$), which gives a distribution with mean 5 days and standard deviation 2 days (18). Supplementary Figure S8 shows that the temporal patterns of inferred infections and reported cases match closely across virtually all countries and age groups. Inferred infections are higher than reported cases for all but a few age groups in a few countries, with larger differences for younger age groups. This is as expected given under-reporting of symptomatic infections, the high proportion of asymptomatic infections, and the decrease in the asymptomatic proportion with increasing age (31).

#### Limitations

The main limitations of our analysis are that it only provides a theoretical upper bound on the potential number of hospitalisations and deaths that could occur in the different countries considered, and that it does not account for waning of immunity, the rollout of boosters or the emergence of new variants. Our approach is also reliant on a number of relatively strong assumptions. Firstly we assume that COVID-19 deaths are completely and sufficiently consistently reported across different countries such that they can be used to estimate and compare cumulative infection burden between countries. However, even among European countries, where reporting of COVID-19 deaths is likely to be more complete than elsewhere, it is possible that there is some under-reporting of COVID-19 deaths, and definitions for COVID-19 deaths differ to some extent between countries and in some countries have changed over time (34). Potential differences in reporting of COVID-19 deaths could be further explored by examining the ratio of excess deaths to COVID-19 deaths reported in each country over time (35).

We also assume that the delay between infection and reported date of death and the age-dependent IFR in unvaccinated individuals are similar enough across countries that we can use the same values for all countries when inferring underlying infections. Different countries use different types of date (occurrence, registration, report) to report deaths so there is likely some variation in the delay between infection and reported death date (34) and there may also be some variation in the delay between infection and occurrence of death between countries due to differences in availability and quality of ICU care. There is also evidence of additional variation in the IFR between countries over that expected just from differences in population age structure (13). However, we believe that the variation in these quantities for the countries considered is likely to be sufficiently small (see Figure 2C in (13) for variation in the population-weighted IFR for the countries we analyse) that it would not significantly change our estimates.

We only consider variation in the IFR due to vaccination, but there are several other potential sources of variation, including differences in severity between variants. Several studies have reported progressively increased severity (corresponding to a higher IFR and/or IHR) for Alpha and Delta in unvaccinated individuals over the original virus strain (36–40), but have not found statistically significant differences in relative risks of hospitalisation and death between variants for vaccinated and unvaccinated individuals (38,39). The severity of different variants may therefore differ less for vaccinated individuals than unvaccinated individuals. Furthermore, comparison of our estimates for the cumulative proportion infected or vaccinated in England with age-stratified seroprevalence estimates suggests that our approach captures the overall change in the IFR that has occurred over time relatively accurately (Supplementary Figure S3).

The approximation of the split of infections between unexposed & unvaccinated individuals and vaccinated individuals that we use to construct the time-varying IFR (Equation (3)) assumes that vaccination rollout happened early and fast enough in most countries that the vaccinated proportion of the population dominates the proportion previously infected. It also assumes that vaccine efficacy against infection is relatively high so the number of breakthrough infections is small relative to the number of infections in unvaccinated individuals, and the current number of vaccinated individuals can be roughly approximated by the vaccination coverage. Whilst these assumptions are reasonable for most of the European countries we consider and for the time period we analyse, they are less valid for other countries where vaccine rollout has been slower, which limits the potential applicability of the analysis beyond the countries considered. The deconvolution method also does not account for reinfections, but to date the proportion of infections that are reinfections appears to be relatively small and protection against severe outcomes from previous infection (20) likely means that the contribution of reinfections to deaths is small.

Another limitation of our analysis is that the deconvolution method relies on sufficient numbers of deaths in each age group to produce stable and temporally accurate estimates of infections. Combined with the approximately exponential increase in the IFR with age, this means that we have to use a large lowest age group (0-39yrs) to obtain reliable infection estimates for countries with low numbers of COVID-19 deaths. This results in a loss of information about differences in infection incidence between age groups <40yrs, especially between adults and children, given many children are still unvaccinated and therefore at higher risk of infection (41). As we assume the same infection risk for all individuals within each age group, the large lowest age group also means that we do not account for variation in infection risk for individuals <40yrs, despite the fact that infection risk varies with age in this group due to heterogeneity in contact patterns (31,42). The estimation issue created by low numbers of deaths in some countries could potentially be resolved by using age-stratified hospitalisation data in addition to, or instead of, death data. However, while this could provide more accurate estimates within each country, such data is not publicly available for all countries and differences in admission criteria between countries would limit comparability.

We assume that immunity acquired through infection provides approximately the same protection against infection, hospitalisation and death as two doses of a viral vector vaccine or mRNA vaccine. This is based on findings of similar levels of protection against symptomatic infection with the Delta variant from prior natural infection and two doses of the AstraZeneca/Pfizer vaccines in a large infection study in the UK (32), and estimates from some studies suggesting higher protection against hospitalisation and death from vaccination than prior infection (at least in individuals aged ≥65) (43) and estimates from others the opposite (44,45). However, some studies have observed higher rates of reinfection among previously infected individuals than vaccinees (46–48), suggesting that we may be underestimating the potential remaining burden among previously infected individuals. On the other hand, we do not account for stronger protection against reinfection among vaccinees who have been previously infected (44,45), which would have the opposite effect on the remaining burden estimates.

#### Code

The code and data underlying this analysis are available online at https://github.com/LloydChapman/covid_remaining_burden. The potential remaining burden estimates can be found in the output folder.

**Supplementary Table S1. Efficacies of 1st and 2nd doses of different vaccines against different COVID-19 outcomes**

| Description | Symbol | Value | Source(s) |
|---|---|---|---|
| **Type A vaccines (AstraZeneca, Janssen, Sputnik, Beijing CNBG)** | | | |
| **“Other” and Alpha variants ($k = 1, 2$)** | | | |
| **Dose 1** | | | |
| Efficacy against infection | $e_{i,a,1,k}$ $(k = 1, 2)$ | 0.70 | (49–51) |
| Efficacy against disease given infection | $e_{d,a,1,k}$ $(k = 1, 2)$ | 0.00 | (50,52,53) |
| Efficacy against hospitalisation given disease | $e_{h,a,1,k}$ $(k = 1, 2)$ | 0.50 | (54–56) |
| Efficacy against death given disease | $e_{m,a,1,k}$ $(k = 1, 2)$ | 0.50 | (56,57) |
| **Dose 2** | | | |
| Efficacy against infection | $e_{i,a,2,k}$ $(k = 1, 2)$ | 0.75 | (39,49,50,52) |
| Efficacy against disease given infection | $e_{d,a,2,k}$ $(k = 1, 2)$ | 0.20 | (50,58) |
| Efficacy against hospitalisation given disease | $e_{h,a,2,k}$ $(k = 1, 2)$ | 0.50 | (55,56) |
| Efficacy against death given disease | $e_{m,a,2,k}$ $(k = 1, 2)$ | 0.75 | (59) |
| **Delta variant** | | | |
| **Dose 1** | | | |
| Efficacy against infection | $e_{i,a,1,3}$ | 0.43 | (53,55) |
| Efficacy against disease given infection | $e_{d,a,1,3}$ | 0.09 | (53,55,56,60,61) |

| | | | |
|---|---|---|---|
| Efficacy against hospitalisation given disease | $e_{h,a,1,3}$ | 0.67 | (56,61,62) |
| Efficacy against death given disease | $e_{m,a,1,3}$ | 0.67 | (56) |
| **Dose 2** | | | |
| Efficacy against infection | $e_{i,a,2,3}$ | 0.63 | (53,55) |
| Efficacy against disease given infection | $e_{d,a,2,3}$ | 0.05 | (53,55,56,60,61) |
| Efficacy against hospitalisation given disease | $e_{h,a,2,3}$ | 0.86 | (56,61,62) |
| Efficacy against death given disease | $e_{m,a,2,3}$ | 0.86 | (56) |
| **Type B vaccines (Pfizer, Moderna)** | | | |
| **“Other” and Alpha variants** | | | |
| **Dose 1** | | | |
| Efficacy against infection | $e_{i,b,1,k}$ $(k = 1, 2)$ | 0.70 | (49–51,53,63) |
| Efficacy against disease given infection | $e_{d,b,1,k}$ $(k = 1, 2)$ | 0.00 | (50,52,53) |
| Efficacy against hospitalisation given disease | $e_{h,b,1,k}$ $(k = 1, 2)$ | 0.5 | (52,54–56,64,65) |
| Efficacy against death given disease | $e_{m,b,1,k}$ $(k = 1, 2)$ | 0.5 | (56,57,65) |
| **Dose 2** | | | |
| Efficacy against infection | $e_{i,b,2,k}$ $(k = 1, 2)$ | 0.85 | (39,49,50,53,63,66) |
| Efficacy against disease given infection | $e_{d,b,2,k}$ $(k = 1, 2)$ | 0.33 | (50,52,53,56,66) |

| | | | |
|---|---|---|---|
| Efficacy against hospitalisation given disease | $e_{h,b,2,k}$ $(k = 1, 2)$ | 0.5 | (55,56,65,66) |
| Efficacy against death given disease | $e_{m,b,2,k}$ $(k = 1, 2)$ | 0.5 | (56,57,65,66) |
| **Delta variant** | | | |
| **Dose 1** | | | |
| Efficacy against infection | $e_{i,b,1,3}$ | 0.62 | (53,55) |
| Efficacy against disease given infection | $e_{d,b,1,3}$ | 0 | (53,55,56,60,61) |
| Efficacy against hospitalisation given disease | $e_{h,b,1,3}$ | 0.79 | (56,61,62) |
| Efficacy against death given disease | $e_{m,b,1,3}$ | 0.79 | (56) |
| **Dose 2** | | | |
| Efficacy against infection | $e_{i,b,2,3}$ | 0.8 | (53,55) |
| Efficacy against disease given infection | $e_{d,b,2,3}$ | 0.05 | (53,55,56,60,61) |
| Efficacy against hospitalisation given disease | $e_{h,b,2,3}$ | 0.84 | (56,61,62) |
| Efficacy against death given disease | $e_{m,b,2,3}$ | 0.84 | (56) |

**Supplementary Table S2. Vaccination status reported by participants of Robert Koch Institute (RKI) COVIMO study on vaccine uptake in Germany, 15th Sep - 18th Oct 2021 (n=3309) (from** (8)**)**

| Age group (yrs) | Percentage vaccinated at least once* (%) |
|---|---|
| 18-29 | 88.2 |
| 30-39 | 81.9 |
| 40-49 | 87.8 |
| 50-59 | 88.9 |
| 60-69 | 93.5 |
| 70-79 | 95.5 |
| 80+ | 96.6 |

*These coverages are much higher than the official vaccination coverages for broader age groups (12-17, 18-59, 60+yrs) reported by RKI (7), likely due to selection bias in the survey participants, but can be used to calculate relative vaccination coverage within the broader age groups.

**Supplementary Table S3. Distributions of durations of infection stages in difference equation model**

| Waiting time | Distribution* | Mean (days) | Reference |
|---|---|---|---|
| Latent period duration, $d_E$ | $Gamma(2.5, 1)$ | 2.5 | (18,67) |
| Subclinical latent period duration, $d_L$ | $Gamma(2.5, 1)$ | 2.5 | (18) |
| Preclinical infection duration, $d_P$ | $Gamma(4, 0.625)$ | 2.5 | (18,67) |
| Clinical infection duration, $d_C$ | $Gamma(4, 0.625)$ | 2.5 | (18) |
| Subclinical infection duration, $d_S$ | $Gamma(4, 1.25)$ | 5 | (18) |

*All Gamma distributions are discretised to be in time steps of 1 day, as the data and difference equation model are discrete with day time steps.

**Supplementary Table S4. Estimates of numbers in different immune states and potential remaining burden of COVID-19 hospitalisations and deaths in 19 European countries as of 21st November 2021***

| Country | Population (millions) | Vaccination coverage (%) | Unvaccinated & unexposed (millions) | Vaccinated & uninfected (millions) | Previously infected (millions) | Proportion unvaccinated & unexposed (%)** | Proportion vaccinated & uninfected (%)** | Proportion previously infected (%)** | Maximum remaining hospitalisations | Maximum remaining deaths | Maximum remaining hospitalisations/ 100,000 people | Maximum remaining deaths/100,000 people |
|---|---|---|---|---|---|---|---|---|---|---|---|---|
| Austria | 9.0 | 65.1 | 2.7 (2.5-2.8) | 5.3 (5.2-5.3) | 1.0 (0.8-1.2) | 30 (28-31) | 59 (57-59) | 11 (9-14) | 63000 (51000-77000) | 11000 (9300-12000) | 700 (570-860) | 120 (100-140) |
| Belgium | 11.6 | 75.0 | 2.2 (1.9-2.4) | 7.3 (7-7.4) | 2.0 (1.7-2.4) | 19 (17-21) | 63 (61-64) | 17 (15-21) | 74000 (57000-92000) | 16000 (14000-19000) | 640 (490-800) | 140 (120-160) |
| Czechia | 10.7 | 57.6 | 3.0 (2.5-3.3) | 4.3 (4.1-4.5) | 3.3 (2.9-3.8) | 28 (23-30) | 40 (38-42) | 31 (27-35) | 79000 (63000-96000) | 14000 (12000-16000) | 740 (590-900) | 130 (110-150) |
| Denmark | 5.8 | 71.8 | 1.5 (1.4-1.6) | 4.0 (3.4-4) | 0.3 (0.2-1.9) | 26 (24-27) | 69 (58-69) | 5 (4-32) | 20000 (16000-28000) | 2900 (2600-3900) | 350 (280-480) | 51 (44-68) |
| England | 56.0 | 73.5 | 10.2 (9.1-10.9) | 30.3 (29.2-31.2) | 15.1 (13.6-16.8) | 18 (16-20) | 54 (52-56) | 27 (24-30) | 180000 (140000-220000) | 33000 (28000-38000) | 320 (250-390) | 58 (50-67) |
| Finland | 5.5 | 76.0 | 1.2 (1.1-1.2) | 4.0 (3.9-4) | 0.4 (0.3-0.6) | 21 (19-22) | 72 (70-73) | 7 (5-10) | 31000 (24000-45000) | 5900 (4300-8900) | 560 (430-820) | 110 (78-160) |
| France | 65.3 | 78.4 | 11.5 (10.8-11.9) | 43.0 (41.9-43.6) | 10.6 (9.7-11.8) | 18 (17-18) | 66 (64-67) | 16 (15-18) | 360000 (280000-460000) | 80000 (68000-94000) | 560 (430-700) | 120 (100-140) |
| Germany | 83.8 | 64.7 | 25.5 (24.6-26) | 48.7 (48.3-49.1) | 9.4 (8.6-10.4) | 30 (29-31) | 58 (58-59) | 11 (10-12) | 820000 (650000-1e+06) | 170000 (150000-190000) | 970 (770-1200) | 200 (170-220) |
| Greece | 10.4 | 62.8 | 2.7 (2.3-2.9) | 5.1 (5-5.3) | 2.4 (2.1-2.8) | 26 (22-28) | 49 (47-51) | 23 (20-26) | 120000 (89000-160 | 30000 (25000-360 | 1200 (850-1500) | 290 (240-350) |

| | | | | | | | | | | | | |
|---|---|---|---|---|---|---|---|---|---|---|---|---|
| | | | | | | | | | 000) | 00) | | |
| Hungary | 9.7 | 60.9 | 0.4 (0.1-0.8) | 1.9 (1.3-2.3) | 7.0 (6.3-7.8) | 5 (1-9) | 19 (13-23) | 72 (65-80) | 43000 (30000-56000) | 11000 (8500-13000) | 450 (310-580) | 110 (88-130) |
| Italy | 60.5 | 76.3 | 11.5 (10.4-11.9) | 38.3 (37.6-38.9) | 10.4 (9.5-11.6) | 19 (17-20) | 63 (62-64) | 17 (16-19) | 370000 (3e+05-440000) | 73000 (64000-81000) | 610 (490-740) | 120 (110-130) |
| Netherlands | 17.1 | 71.6 | 4.4 (4.2-4.5) | 11.4 (11.3-11.5) | 1.2 (1-1.5) | 26 (25-26) | 67 (66-67) | 7 (6-9) | 180000 (140000-220000) | 40000 (34000-46000) | 1000 (800-1300) | 230 (200-270) |
| Norway | 5.4 | 77.5 | 1.2 (1.1-1.2) | 4.1 (4-4.1) | 0.1 (0.1-0.3) | 22 (21-22) | 75 (74-76) | 3 (2-5) | 20000 (16000-26000) | 3300 (2700-4000) | 380 (300-470) | 61 (50-74) |
| Portugal | 10.2 | 87.6 | 1.1 (0.9-1.1) | 7.6 (7.3-7.7) | 1.5 (1.3-1.9) | 10 (9-11) | 74 (71-76) | 15 (13-18) | 27000 (22000-33000) | 5100 (4500-5900) | 270 (210-320) | 50 (44-58) |
| Romania | 19.2 | 33.5 | 2.0 (1.5-3.2) | 2.3 (1.6-2.9) | 14.3 (12.8-15.5) | 11 (8-16) | 12 (8-15) | 74 (66-80) | 380000 (260000-5e+05) | 1e+05 (87000-120000) | 2000 (1400-2600) | 540 (450-630) |
| Slovakia | 5.5 | 44.3 | 1.3 (0.9-1.6) | 1.4 (1.3-1.5) | 2.6 (2.3-3.2) | 25 (16-29) | 25 (23-27) | 48 (43-58) | 53000 (41000-66000) | 11000 (9500-13000) | 970 (760-1200) | 210 (170-240) |
| Slovenia | 2.1 | 56.6 | 0.8 (0.7-0.8) | 1.0 (1.0-1.0) | 0.3 (0.2-0.4) | 37 (33-39) | 48 (46-49) | 15 (12-20) | 23000 (18000-29000) | 4300 (3700-5100) | 1100 (870-1400) | 210 (180-250) |
| Spain | 46.8 | 81.3 | 6.7 (6.1-7.1) | 31.1 (30.3-31.6) | 8.8 (8-9.9) | 14 (13-15) | 66 (65-68) | 19 (17-21) | 150000 (120000-180000) | 24000 (21000-28000) | 310 (250-380) | 52 (45-59) |
| Sweden | 10.1 | 69.1 | 2.4 (2.1-2.6) | 6.0 (5.8-6.1) | 1.7 (1.4-2.1) | 24 (21-25) | 59 (57-61) | 16 (14-21) | 50000 (40000-61000) | 9000 (7600-11000) | 500 (390-600) | 89 (75-110) |

*Numbers in parentheses are 95% credible intervals. **Proportions sum to slightly less than 100% as currently infected individuals are not included.

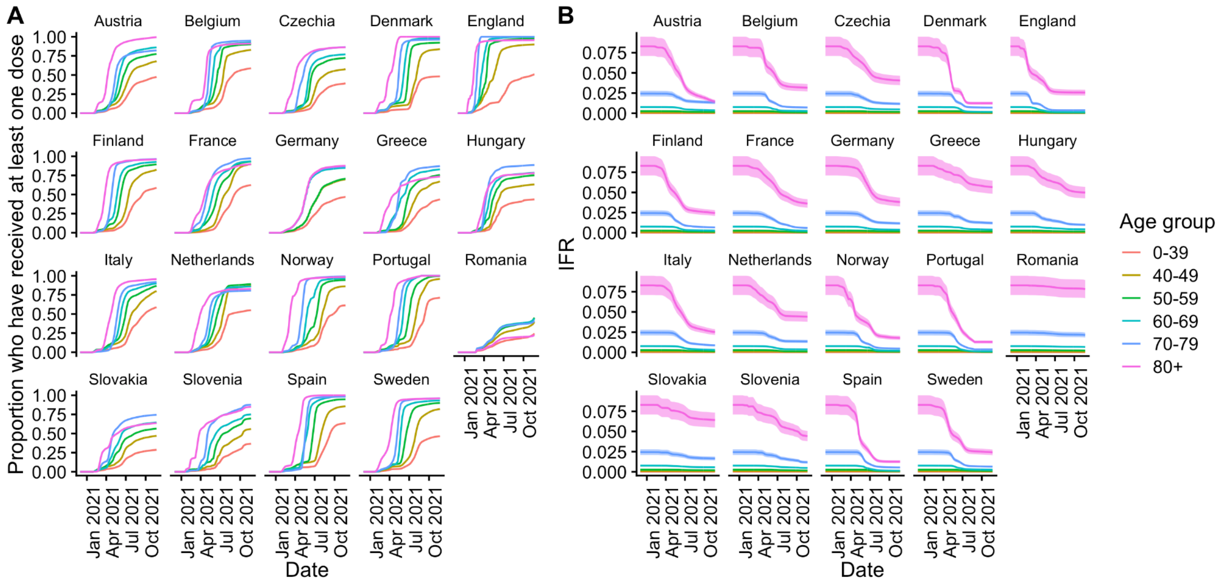


**Supplementary Figure S1. (A) Vaccine coverage and (B) IFR over time (calculated using Equation (3)) by age group and country since 1st December 2020.** Shaded bands in (B) show 95% credible intervals based on a truncated normal approximation to posterior distribution of IFR in (13).

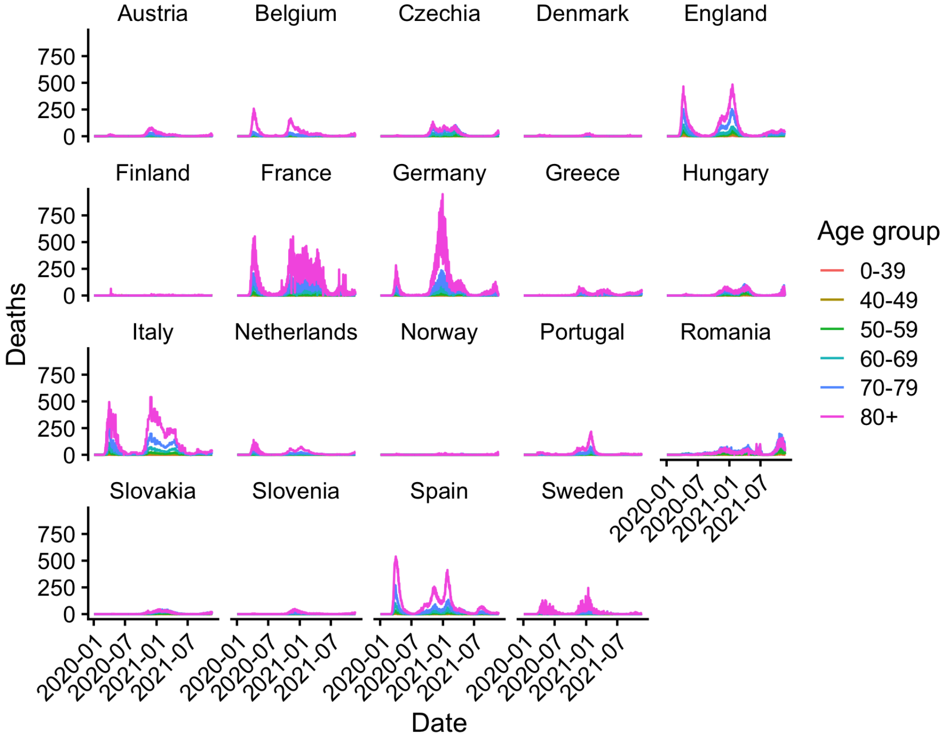


**Supplementary Figure S2. Age-stratified death time series from the COVerAGE database** (1,2) **used to infer age-stratified infection time series in Figure 2A in the main text.**

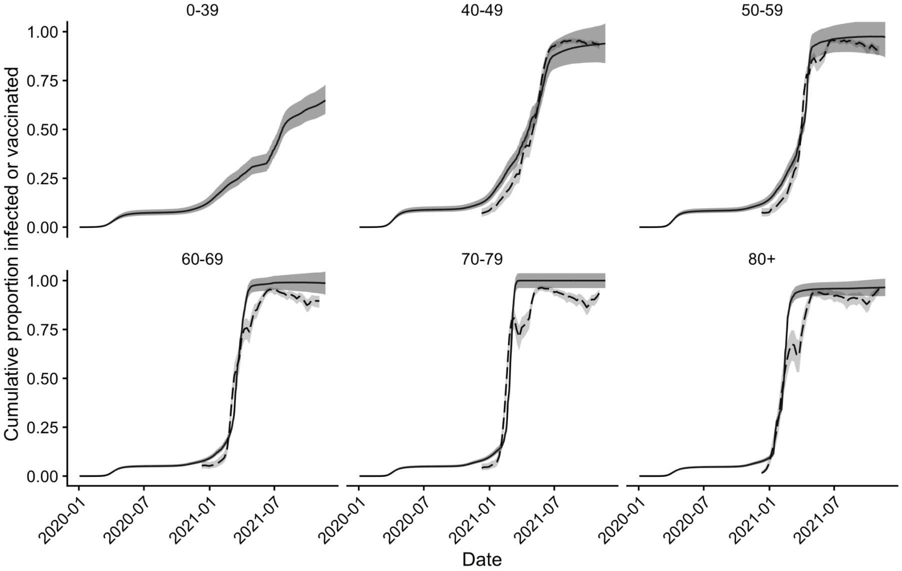


**Supplementary Figure S3. Estimated cumulative proportion infected or vaccinated in England (solid lines) and modelled seroprevalence estimates for England (dashed lines) from the UK Office of National Statistics (ONS)** (68)**.** Shaded regions show 95% credible intervals. Seroprevalence estimates for 0-39-year-olds are not available as the serological testing in the ONS Coronavirus Infection Survey only covers adults aged ≥16yrs.

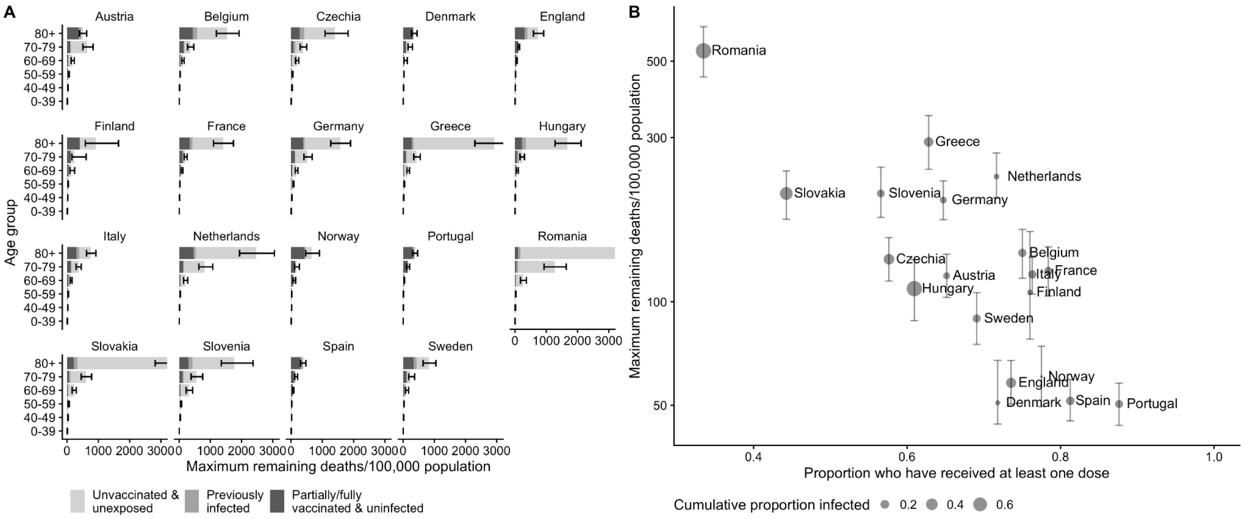


**Supplementary Figure S4. (A) Maximum remaining COVID-19 deaths by age group and country (assuming no waning of immunity or emergence of immune escape variants), and (B) relationship between overall remaining deaths per 100,000 population and proportion who have received at least one vaccine dose across countries.** Note upper limit of horizontal axis in (A) has been truncated to ensure differences between countries remain visible and vertical axis in (B) is on log scale. Supplementary Figure S6B shows plot (A) without truncation of the horizontal axis.

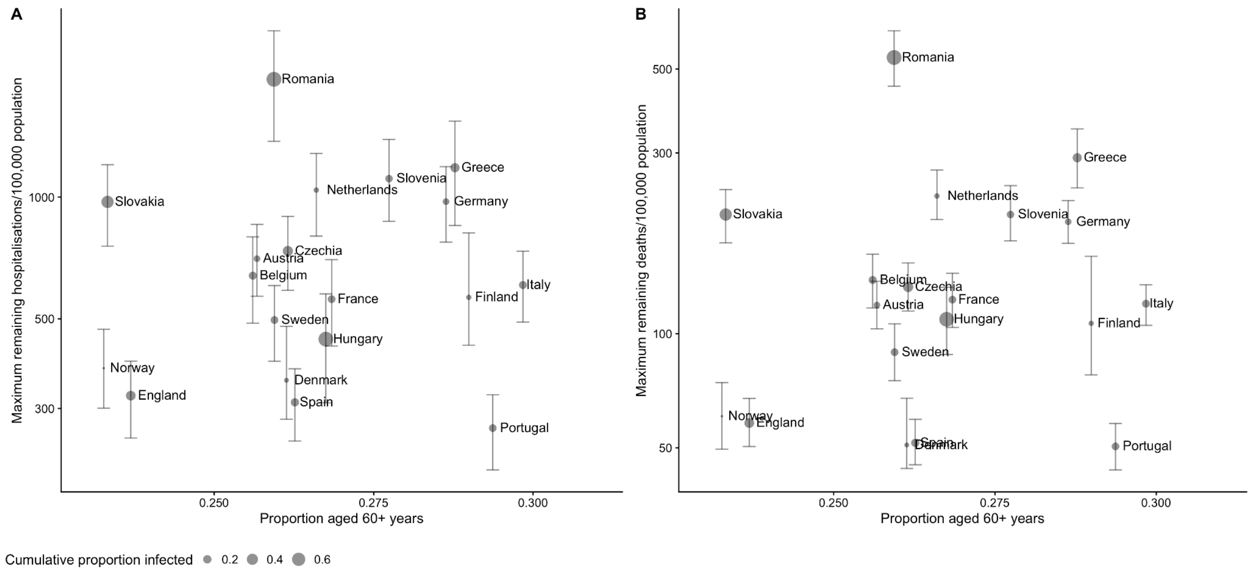


**Supplementary Figure S5. Relationship between remaining burden of (A) hospitalisations and (B) deaths and proportion of the population 60 or more years of age across countries.** Note vertical axes are on the log scale.

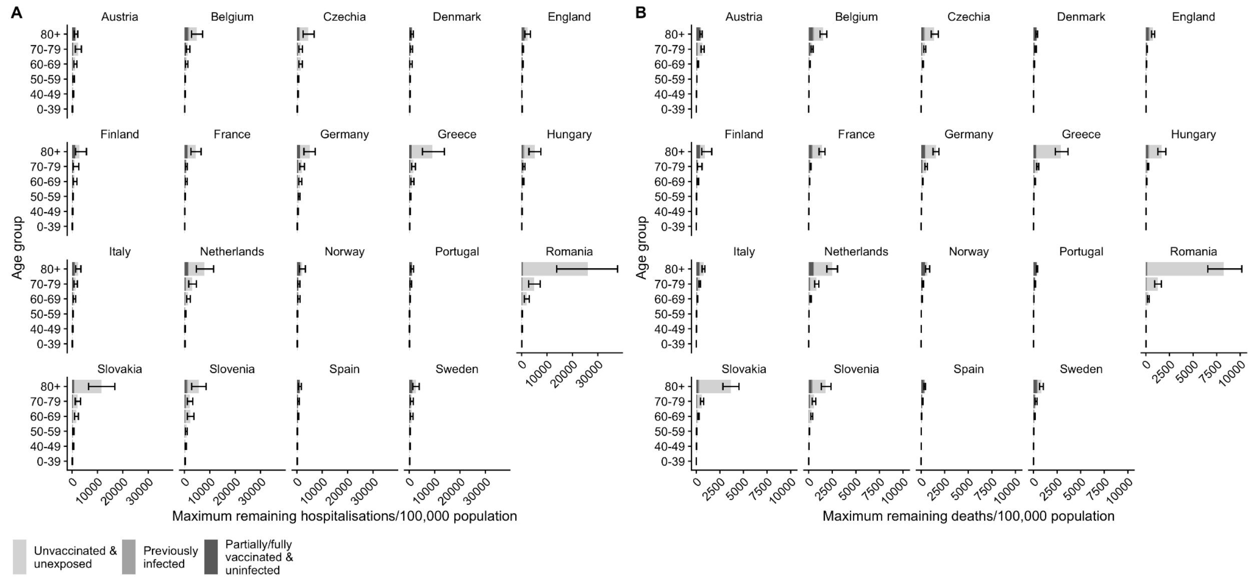


**Supplementary Figure S6. Estimated maximum remaining (A) hospitalisations and (B) deaths per 100,000 population by country, age group and immune status.**

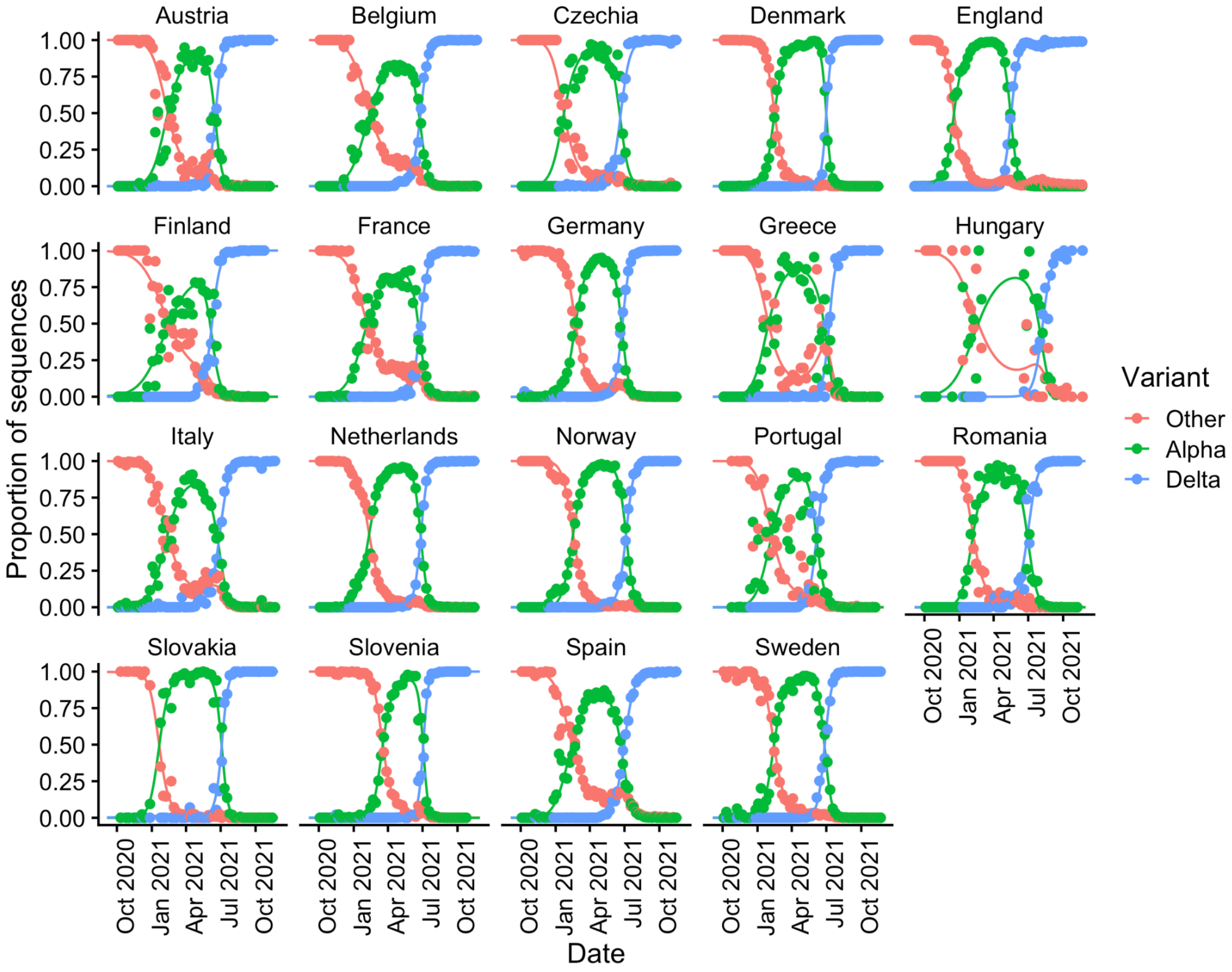


**Supplementary Figure S7. Estimated proportions of different variants over time from multinomial regression model fitted to GISAID (EU countries) and COVID-19 Genomics UK consortium (England) data on numbers of sequences of different variants**

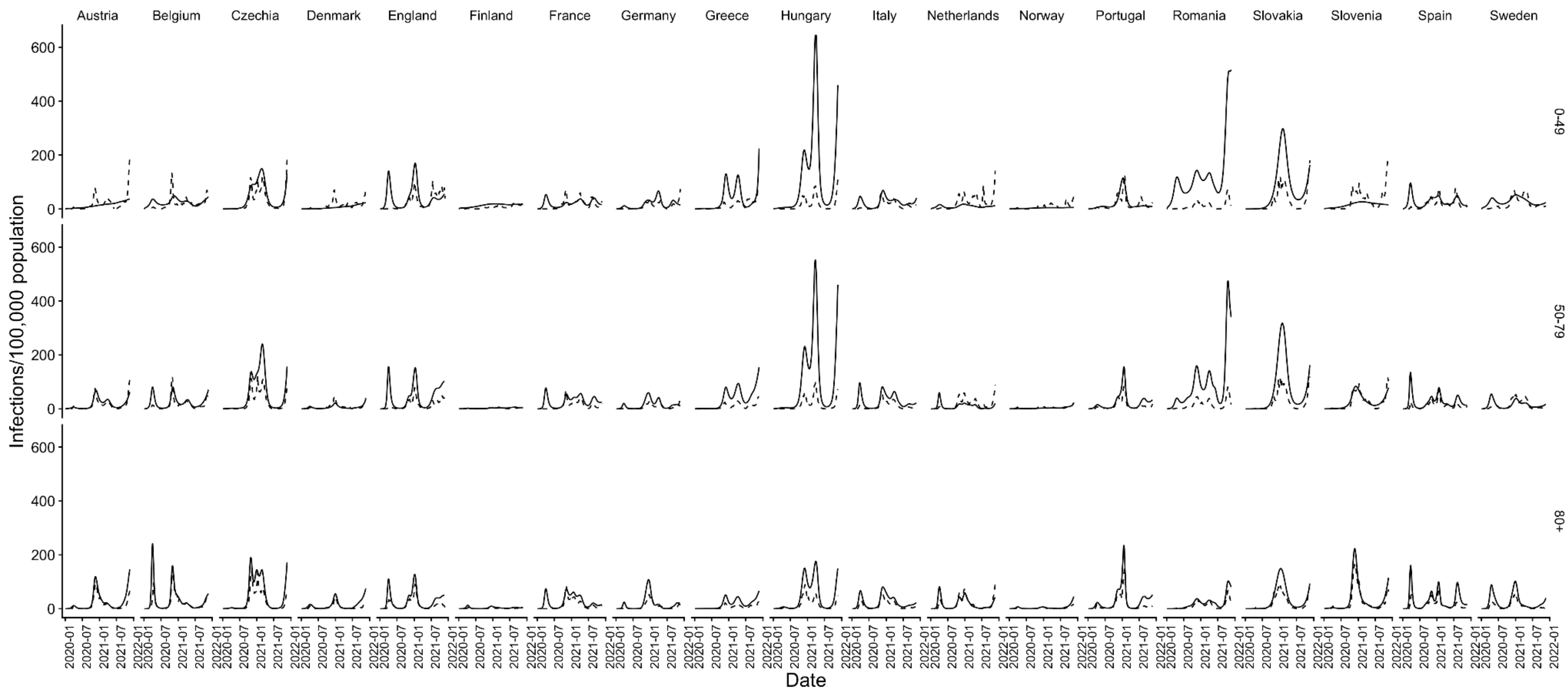


**Supplementary Figure S8. Inferred infection time series (solid lines) and reported cases (dashed lines) by country (columns) and age group (rows).** Reported cases by age group obtained from ECDC database (69) and UK government's COVID-19 dashboard (6), and observed and inferred counts re-aggregated into matching age groups.

28. Comas-Herrera A, Zalakaín J, Lemmon E, Henderson D, Litwin C, Hsu AT, et al. Mortality

associated with COVID-19 in care homes: international evidence [Internet]. International Long-Term Care Policy Network; 2021 Feb [cited 2021 Oct 21]. Available from: https://ltccovid.org/wp-content/uploads/2021/02/LTC_COVID_19_international_report_January-1-February-1-1.pdf

43. Young-Xu Y, Smith J, Korves C. SARS-Cov-2 Infection versus Vaccine-Induced Immunity among Veterans. medRxiv. 2021 Sep 29;2021.09.27.21264194.

62. Stowe J, Andrews N, Gower C, Gallagher E, Utsi L, Simmons R, et al. Effectiveness of COVID-19 vaccines against hospital admission with the Delta (B.1617.2) variant [Internet].

[cited 2021 Oct 22]. Available from: https://media.tghn.org/articles/Effectiveness_of_COVID-19_vaccines_against_hospital_admission_with_the_Delta_B._G6gnnqJ.pdf
